# Structured Hypertension Education and Telephone Follow-up After Emergency Department Discharge: A Prospective Quality Improvement Initiative in Pakistan

**DOI:** 10.64898/2026.09.07.26361893

**Authors:** Mubarra Aqeel, Muhammad Sarmad Waqar Hashmi, Uswah Shoaib, Zarwa Rashid, Ali Shan Hafeez

## Abstract

**Background:** Emergency department (ED) encounters may identify uncontrolled hypertension, but continuity after discharge is often weak. We evaluated a low-cost quality-improvement strategy combining standardized discharge education with early telephone follow-up in a resource-constrained setting.

**Methods:** This prospective, single-center pre-post quality improvement initiative enrolled 50 consecutive adults with diagnosed hypertension or elevated blood pressure at presentation who were clinically stable for discharge from the ED of Shaikh Zayed Hospital, Lahore, Pakistan, during August 2026. Participants received structured counseling on medication adherence, lifestyle measures, blood pressure (BP) monitoring, warning symptoms, and primary-care follow-up, followed by a telephone assessment approximately 2 weeks later. The primary estimand was mean within-participant systolic BP (SBP) reduction among participants who were reached and had complete numeric follow-up SBP and diastolic BP (DBP). We report bootstrap 95% confidence intervals (CIs), robust paired analyses, responder outcomes, and missing-data sensitivity analyses.

**Results:** Thirty-eight of 50 participants (76.0%) were reached and had complete paired BP measurements. In this paired cohort, mean SBP decreased from 164.4 (SD 20.3) to 132.6 (SD 12.4) mmHg, a mean reduction of 31.8 mmHg (95% bootstrap CI 24.4 to 39.3; P<0.001). Mean DBP decreased from 94.7 (SD 8.3) to 84.3 (SD 7.5) mmHg, a mean reduction of 10.4 mmHg (95% bootstrap CI 6.7 to 13.9; P<0.001). Strict BP control (<140/<90 mmHg) was achieved by 20/38 participants (52.6%; exact 95% CI 35.8% to 69.0%). SBP reduction of at least 10 mmHg occurred in 33/38 (86.8%), and DBP reduction of at least 5 mmHg in 26/38 (68.4%). Among reached participants with nonmissing responses, 73.0% reported medication adherence, 81.6% lifestyle modification, 77.1% primary-care follow-up, and 81.1% symptom improvement. Missing-data sensitivity analyses remained directionally consistent with lower follow-up BP.

**Conclusions:** Structured discharge education followed by early telephone contact was associated with substantial short-term within-participant BP reductions and high reported engagement with recommended care. Because the project lacked a concurrent control group, used a short follow-up interval, and had potential selection and measurement biases, the findings should not be interpreted as a causal treatment effect. Controlled evaluation with standardized BP measurement and longer follow-up is warranted.

## Introduction

Hypertension is a major modifiable contributor to cardiovascular, cerebrovascular, and kidney disease. Effective long-term control requires pharmacological treatment when indicated, sustained lifestyle modification, and reliable follow-up; however, achieving and maintaining control remains difficult in many settings. [1,2]

Pakistan carries a substantial hypertension burden, and published work has highlighted gaps in awareness, treatment, adherence, and control.[3-5] These challenges are especially relevant after acute-care encounters, where patients may leave the emergency department (ED) with markedly elevated blood pressure but without an established mechanism for reinforcement of education, reassessment, and linkage to longitudinal care.

The American Heart Association has emphasized that elevated BP is common in acute-care settings and that improved transitions to outpatient management are an important area for practice improvement and research.[6] Quality-improvement work from other resource-limited settings suggests that organized follow-up systems can improve continuity for people with hypertension, while longitudinal data support the importance of regular follow-up for BP control. [7,8]

We therefore implemented a prospective quality improvement initiative in the ED of Shaikh Zayed Hospital, Lahore, using standardized hypertension education before discharge and structured telephone follow-up approximately 2 weeks later. The primary aim was to estimate short-term within-participant change in SBP; secondary aims were to describe DBP change, BP-control and responder outcomes, follow-up feasibility, and patient-reported medication adherence, lifestyle change, primary-care follow-up, and symptom improvement.

## Methods

### Context, setting, and participants

This prospective, single-center, pre-post quality improvement initiative was conducted in the Emergency Department of Shaikh Zayed Hospital, Lahore, Pakistan, during August 2026. The project used a Plan-Do-Study-Act (PDSA) framework to address locally identified gaps in hypertension education and continuity after ED discharge. Reporting was organized with reference to SQUIRE 2.0.[15]

Fifty consecutive adults with diagnosed hypertension or elevated BP at presentation were enrolled using non-probability consecutive sampling if they were clinically stable for discharge, able to communicate by telephone, and willing to participate. Patients with hypertensive emergencies requiring admission, critical illness, pregnancy-related hypertension, inpatient admission for another reason, or inability to participate in telephone follow-up were excluded. Verbal informed consent was obtained before enrollment.

### Intervention

Before discharge, participants received standardized verbal education about hypertension and its complications. Counseling covered adherence to prescribed antihypertensive medication, dietary sodium restriction and other healthy dietary practices, physical activity, weight management, smoking cessation when applicable, home or community BP monitoring, recognition of warning symptoms, and the importance of primary-care follow-up. No written educational leaflet was used. The standardized discharge education was delivered by a physician member of the project team.

Approximately 2 weeks after discharge, participants were contacted by telephone using a structured assessment. They were asked about a recent BP measurement, medication adherence, lifestyle changes, attendance at primary-care follow-up, and symptom change. Follow-up BP could be obtained using an available home BP monitor or at a local clinic or hospital. Participants who could not initially be contacted were recontacted when feasible. The structured follow-up telephone assessments were conducted by a physician member of the project team.

### Outcome measures

The primary outcome was change in SBP from baseline to follow-up among participants with complete paired BP measurements who were documented as reached by telephone. DBP change was a key secondary continuous outcome. Strict BP control was defined as follow-up SBP <140 mmHg and DBP <90 mmHg; an inclusive threshold sensitivity analysis used SBP <=140 mmHg and DBP <=90 mmHg. Supportive responder outcomes were reductions of at least 10 mmHg in SBP and at least 5 mmHg in DBP. These responder thresholds were descriptive and did not replace the continuous primary estimand.

Process and patient-reported outcomes included successful telephone follow-up, acquisition of a complete numeric BP pair, medication adherence, lifestyle modification, primary-care follow-up, and symptom improvement. For patient-reported outcomes, observed proportions used reached participants with nonmissing responses as the denominator; all-enrolled lower-bound analyses were treated as sensitivity summaries rather than observed response rates.

### Data integrity and statistical analysis

Before final analysis, the cleaned dataset underwent an internal consistency audit. The final analytic file contained 50 unique patient identifiers, no duplicates, 38 complete follow-up SBP/DBP pairs, and no discrepancies between numeric BP values and recalculated BP-control, BP-improvement, or BP-reduction variables. The primary paired cohort consisted of the 38 participants documented as reached who had both numeric follow-up SBP and DBP.

Continuous variables were summarized using means and standard deviations (SDs), and categorical variables using counts and percentages. The primary analysis estimated mean within-participant BP reduction (baseline minus follow-up) with 10,000-sample nonparametric bootstrap 95% CIs. Paired t tests, Wilcoxon signed-rank tests, 50,000-draw sign-flip permutation tests, 20% trimmed means, and standardized paired effect sizes (Cohen dz and Hedges gz) were used as robustness analyses. Exact binomial CIs were used for proportions. Baseline differences between the paired cohort and participants without paired follow-up BP were described with standardized mean differences (SMDs), emphasizing magnitude rather than significance testing.

Missing-data robustness analyses included (1) an all-enrolled no-change assumption in which baseline BP was carried forward for participants without paired follow-up measurements; (2) multiple imputation under a missing-at-random assumption using predictive mean matching with 50 imputations and baseline SBP, baseline DBP, age, and sex as predictors; (3) delta-adjusted missing-not-at-random scenarios that increased imputed follow-up BP by prespecified amounts; and (4) deterministic tipping-point calculations. All tests were two-sided. Analyses were performed in R using the final locked dataset; reproducibility information and session details were retained with the analysis outputs.

### Ethical considerations

Ethical approval for this quality improvement project was granted by the Technical and Ethical Review Committee (TERC), a review wing of the Institutional Review and Research Advisory Board (IRRAB), Shaikh Zayed Medical Complex, Lahore, Pakistan (TERC ID: TERC/SC/INT/2026/111; approval date: 28 July 2026). Verbal informed consent was obtained from all participants before enrollment.

### Patient and Public Involvement

Patients and members of the public were not involved in the design, conduct, reporting, or dissemination plans of this quality improvement initiative. Patients participated in the initiative as recipients of the intervention and in follow-up data collection, but they were not involved as research partners.

## Results

### Participants and baseline characteristics

Fifty participants were enrolled. Thirty-eight (76.0%; exact 95% CI 61.8% to 86.9%) were documented as reached by telephone and had complete numeric follow-up SBP and DBP; these 38 participants comprised the primary paired cohort (Figure 1). Twelve participants did not contribute paired follow-up BP measurements. The full cohort had a mean age of 51.5 years (SD 12.6), 32/50 (64.0%) were female, mean baseline SBP was 165.6 mmHg (SD 19.3), and mean baseline DBP was 94.9 mmHg (SD 7.9). In the paired cohort, mean age was 52.3 years (SD 12.4), 22/38 (57.9%) were female, baseline SBP was 164.4 mmHg (SD 20.3), and baseline DBP was 94.7 mmHg (SD 8.3) (Table 1).

**Figure 1.**
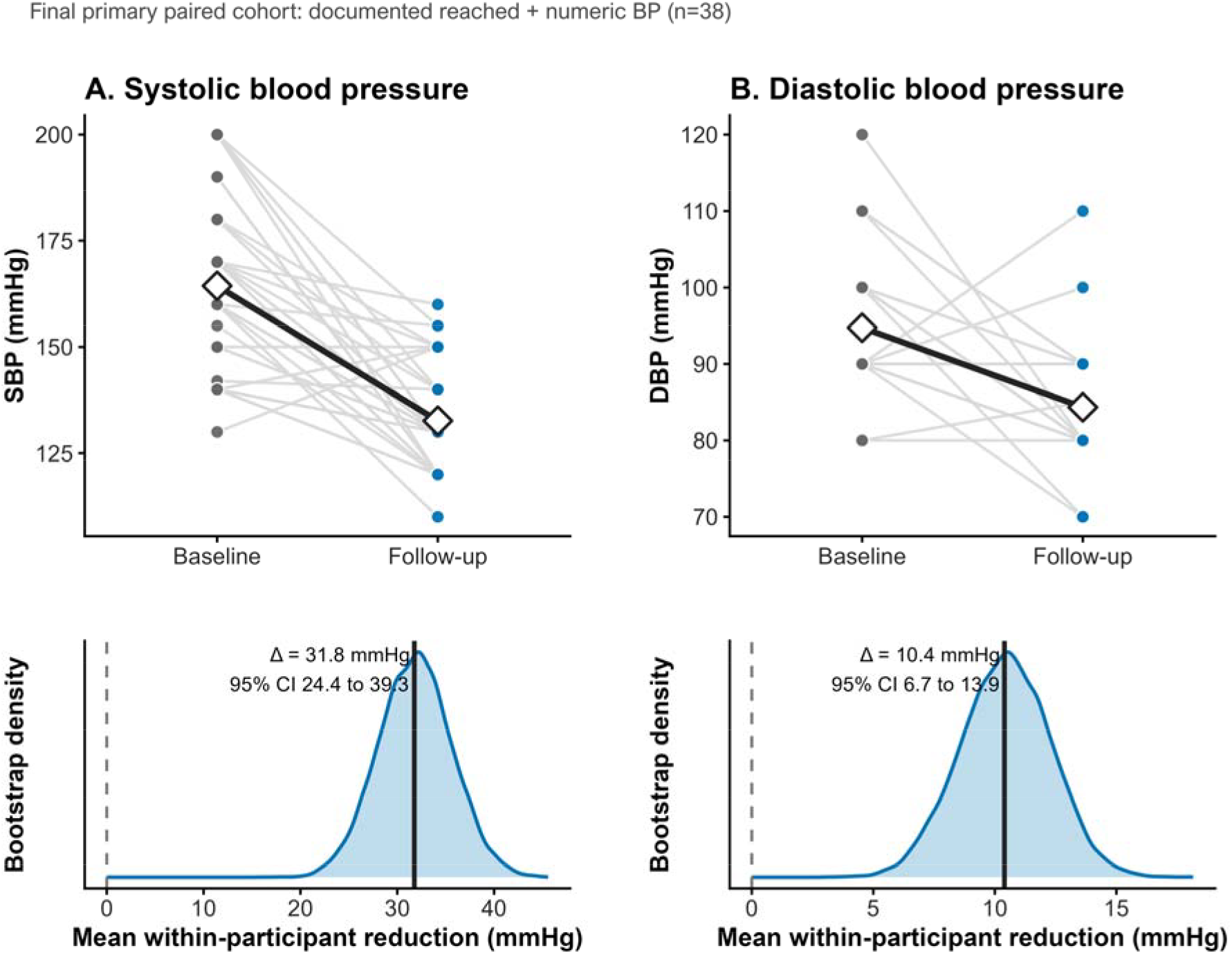
Patient-level blood pressure change with bootstrap estimation. The upper panels show paired baseline-to-follow-up trajectories for systolic and diastolic blood pressure in the final primary cohort (n=38). Thin lines represent individual participants and diamonds represent group means. The lower panels show the bootstrap distribution of the mean within-participant reduction; vertical solid lines show the observed mean reduction and dashed lines denote zero change. Positive reductions indicate lower blood pressure at follow-up.

**Table 1.**
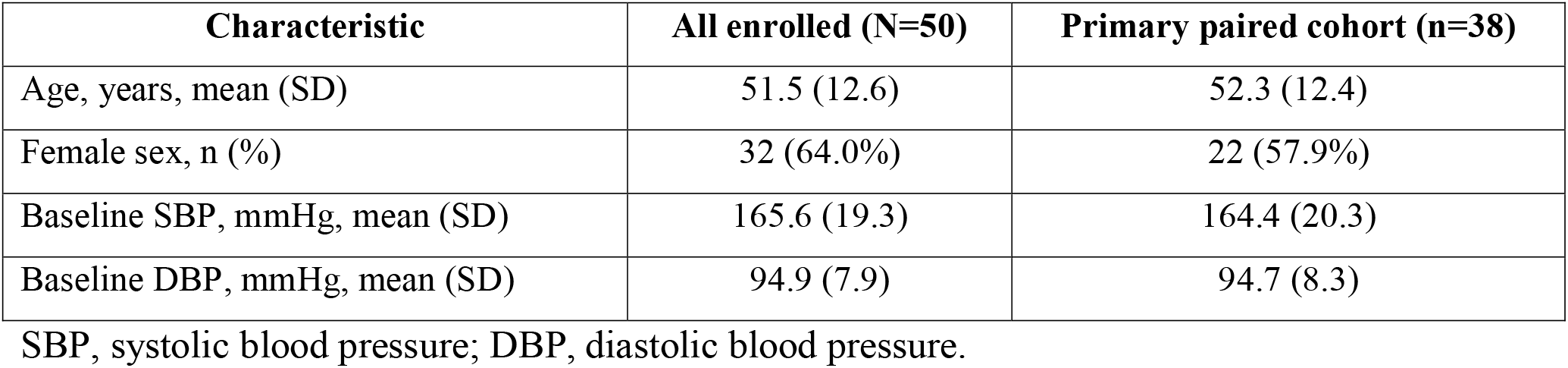
Baseline characteristics of the enrolled cohort and primary paired cohort.

Baseline imbalance between participants with and without paired follow-up data was most pronounced for sex (absolute SMD 0.58), with smaller imbalances for baseline SBP (0.29), age (0.24), and baseline DBP (0.10). These differences indicate potential selection or attrition bias and were considered in interpretation (Supplementary Figure S1).

### Blood pressure outcomes

Among the 38 participants with paired measurements, mean SBP decreased from 164.4 mmHg (SD 20.3) at baseline to 132.6 mmHg (SD 12.4) at follow-up, corresponding to a mean within-participant reduction of 31.8 mmHg (95% bootstrap CI 24.4 to 39.3; paired t test P<0.001; Wilcoxon P<0.001; Cohen dz=1.35). Mean DBP decreased from 94.7 mmHg (SD 8.3) to 84.3 mmHg (SD 7.5), a mean reduction of 10.4 mmHg (95% bootstrap CI 6.7 to 13.9; paired t test P<0.001; Wilcoxon P<0.001; Cohen dz=0.88) (Table 2; Figure 1). Sign-flip permutation tests were also significant for both outcomes (P<0.001).

**Table 2.**
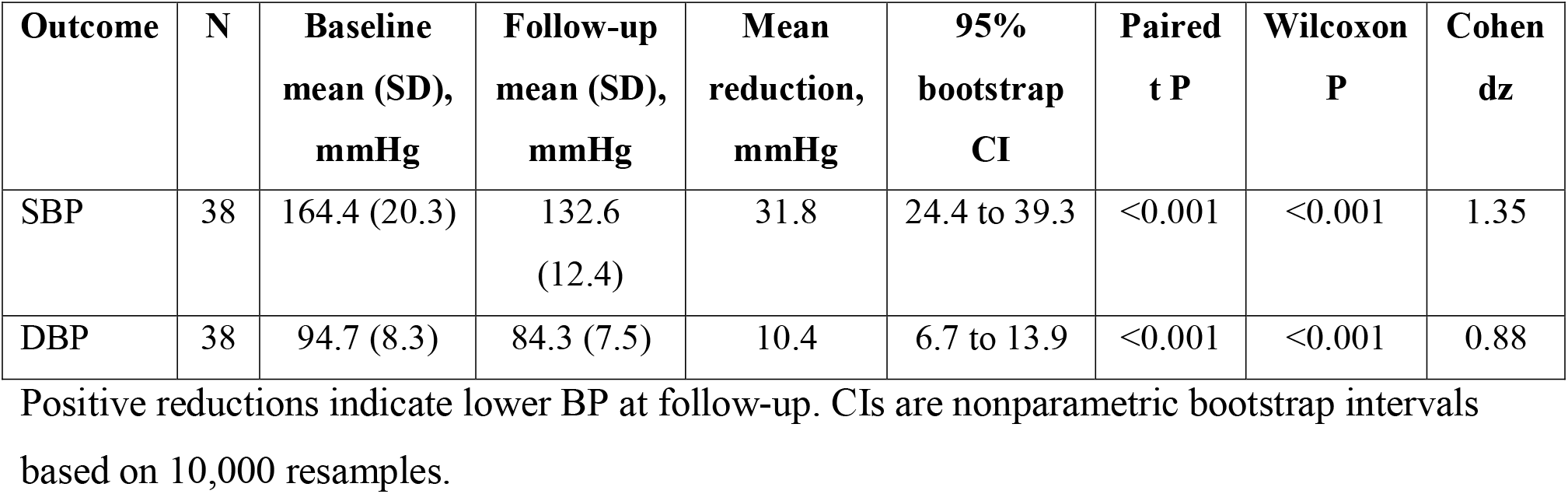
Primary paired blood pressure outcomes.

| **Outcome** | **N** | **Baseline**<br>**mean (SD),**<br>**mmHg** | **Follow-up**<br>**mean (SD),**<br>**mmHg** | **Mean**<br>**reduction,**<br>**mmHg** | **95%**<br>**bootstrap**<br>**CI** | **Paired**<br>**t P** | **Wilcoxon**<br>**P** | **Cohen**<br>**dz** |
| --- | --- | --- | --- | --- | --- | --- | --- | --- |
| SBP | 38 | 164.4 (20.3) | 132.6<br>(12.4) | 31.8 | 24.4 to 39.3 | <0.001 | <0.001 | 1.35 |
| DBP | 38 | 94.7 (8.3) | 84.3 (7.5) | 10.4 | 6.7 to 13.9 | <0.001 | <0.001 | 0.88 |

Positive reductions indicate lower BP at follow-up. CIs are nonparametric bootstrap intervals based on 10,000 resamples.

Individual trajectories were heterogeneous but predominantly favorable. SBP improved in 35/38 participants (92.1%), was unchanged in 1/38 (2.6%), and worsened in 2/38 (5.3%). DBP improved in 26/38 (68.4%), was unchanged in 9/38 (23.7%), and worsened in 3/38 (7.9%). Joint SBP/DBP transitions are shown in Figure 2, and the distribution of individual response magnitudes in Figure 3.

**Figure 2.**
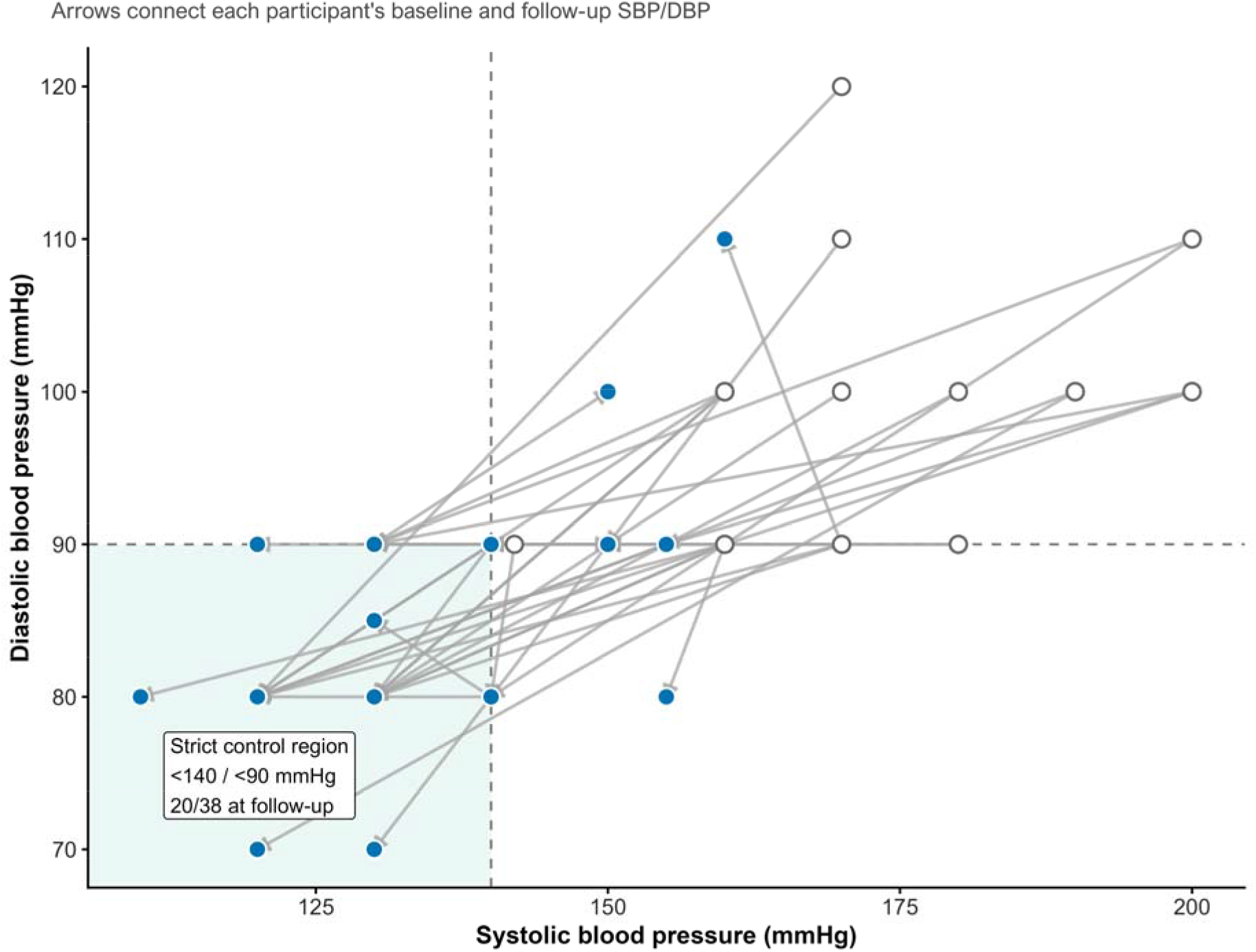
Joint blood pressure state transitions after ED discharge. Joint systolic and diastolic blood pressure state transitions after emergency-department discharge. Each arrow connects an individual participant’s baseline SBP/DBP to the corresponding follow-up measurement. The shaded lower-left region represents the prespecified strict control definition of SBP <140 mmHg and DBP <90 mmHg. Baseline values are open points and follow-up values are filled points.

**Figure 3.**
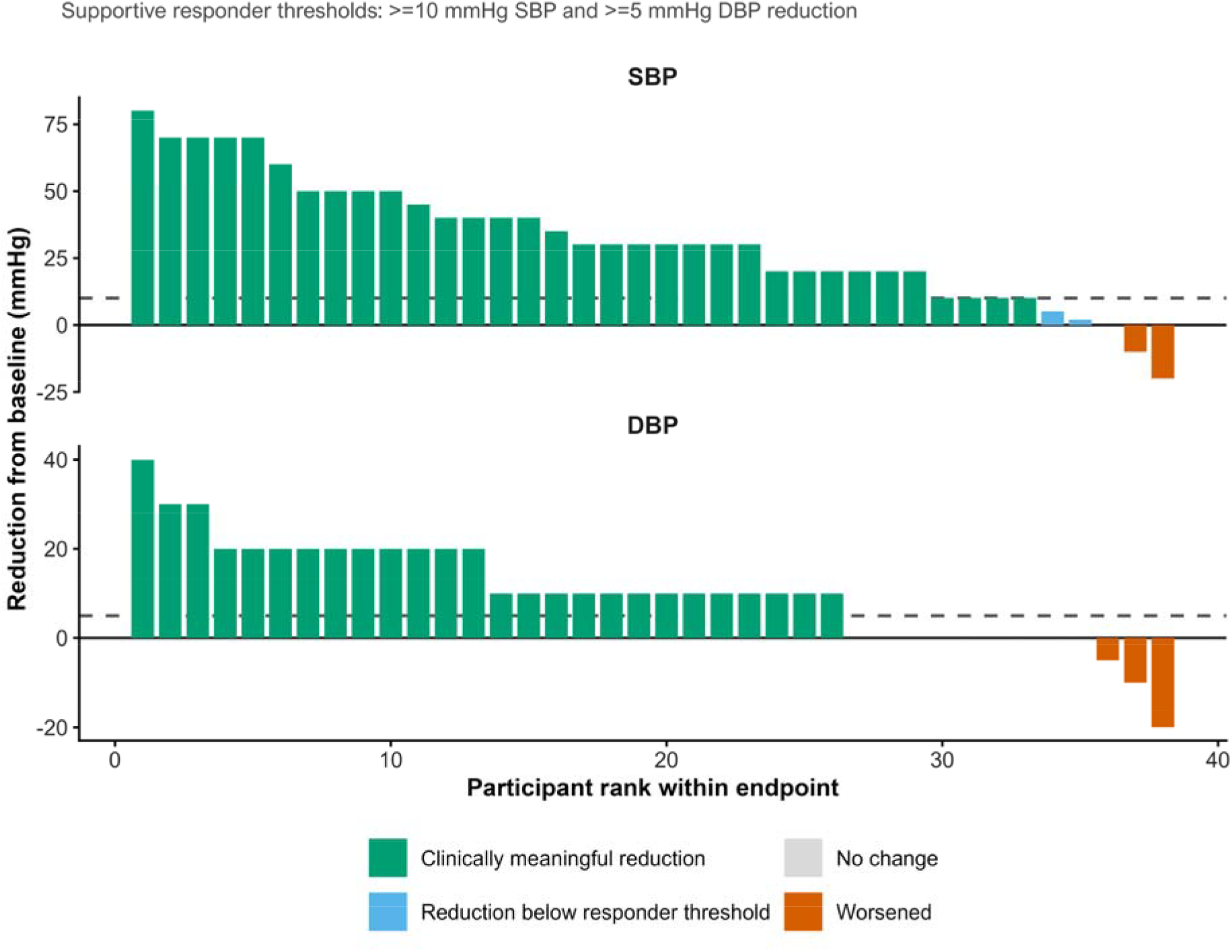
Heterogeneity of individual blood pressure response. Participants are ranked within endpoint by within-participant reduction. Positive bars indicate lower BP at follow-up and negative bars indicate worsening. Dashed horizontal lines denote supportive responder thresholds of at least 10 mmHg SBP reduction and at least 5 mmHg DBP reduction. These are descriptive secondary thresholds and do not replace the continuous primary estimand.

### Blood pressure control and responder outcomes

Strict BP control (SBP <140 mmHg and DBP <90 mmHg) was achieved by 20/38 participants (52.6%; exact 95% CI 35.8% to 69.0%). Under the inclusive <=140/<=90 mmHg sensitivity definition, 30/38 (78.9%; 95% CI 62.7% to 90.4%) met the threshold. Thirty-three of 38 participants (86.8%; 95% CI 71.9% to 95.6%) achieved an SBP reduction of at least 10 mmHg, and 26/38 (68.4%; 95% CI 51.3% to 82.5%) achieved a DBP reduction of at least 5 mmHg (Table 3).

**Table 3.** Clinical, process, and patient-reported follow-up outcomes.

| **Domain** | **Outcome** | **Events/N** | **%** | **Exact 95%**<br>**CI** |
| --- | --- | --- | --- | --- |
| Process | Telephone follow-up reached | 38/50 | 76.0 | 61.8 to 86.9 |
| Process | Reached + complete numeric BP pair | 38/50 | 76.0 | 61.8 to 86.9 |
| Clinical | Strict BP control: SBP <140 and DBP <90 | 20/38 | 52.6 | 35.8 to 69.0 |
| Clinical | Threshold sensitivity: SBP ≤140 and DBP ≤90 | 30/38 | 78.9 | 62.7 to 90.4 |
| Clinical | SBP reduction ≥10 mmHg | 33/38 | 86.8 | 71.9 to 95.6 |
| Clinical | DBP reduction ≥5 mmHg | 26/38 | 68.4 | 51.3 to 82.5 |
| Clinical | Any BP improvement (SBP or DBP lower) | 35/38 | 92.1 | 78.6 to 98.3 |
| Patient-reported | Medication adherence | 27/37 | 73.0 | 55.9 to 86.2 |
| Patient-reported | Lifestyle modification | 31/38 | 81.6 | 65.7 to 92.3 |
| Patient-reported | Primary-care follow-up | 27/35 | 77.1 | 59.9 to 89.6 |
| Patient-reported | Symptoms improved | 30/37 | 81.1 | 64.8 to 92.0 |

Patient-reported outcomes use reached participants with nonmissing responses as the observed denominator. BP, blood pressure; SBP, systolic blood pressure; DBP, diastolic blood pressure.

### Follow-up feasibility and patient-reported outcomes

Telephone follow-up with a complete numeric BP pair was obtained for 38/50 participants (76.0%; 95% CI 61.8% to 86.9%). Among reached participants with nonmissing responses, medication adherence was reported by 27/37 (73.0%; 95% CI 55.9% to 86.2%), lifestyle modification by 31/38 (81.6%; 95% CI 65.7% to 92.3%), primary-care follow-up by 27/35 (77.1%; 95% CI 59.9% to 89.6%), and symptom improvement by 30/37 (81.1%; 95% CI 64.8% to 92.0%) (Table 3; Figure 5). Conservative all-enrolled lower-bound estimates are provided in the Supplementary Material.

### Robustness and missing-data sensitivity analyses

The estimated reductions remained positive across sensitivity analyses (Figure 4; Supplementary Tables S1-S2). Under an all-enrolled no-change assumption for participants without paired follow-up BP, the mean reduction was 24.1 mmHg for SBP (95% bootstrap CI 17.6 to 30.8) and 7.9 mmHg for DBP (95% bootstrap CI 4.9 to 11.0). Multiple imputation under a missing-at-random assumption estimated reductions of 32.6 mmHg (95% CI 26.2 to 39.1) for SBP and 10.2 mmHg (95% CI 6.7 to 13.7) for DBP. Directionally favorable estimates also persisted under prespecified adverse delta-adjusted missing-not-at-random scenarios.

**Figure 4.**
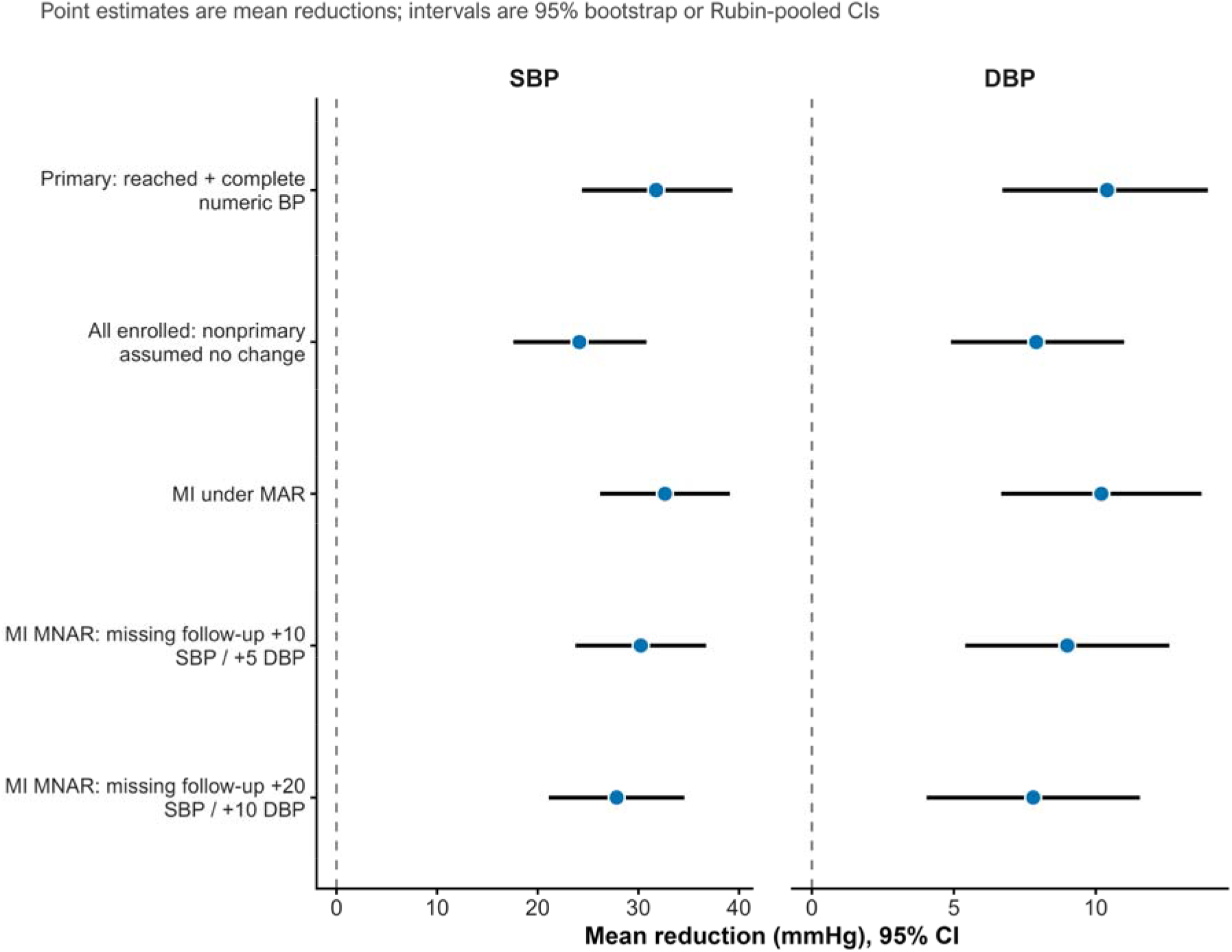
Robustness of estimated blood pressure reduction to missing-data assumptions. Points show estimated mean reductions and horizontal lines show 95% bootstrap or Rubin-pooled confidence intervals. Analyses include the final primary paired cohort, an all-enrolled no-change assumption, multiple imputation under missing at random, and adverse delta-adjusted missing-not-at-random scenarios. Positive values indicate lower follow-up blood pressure.

**Figure 5.**
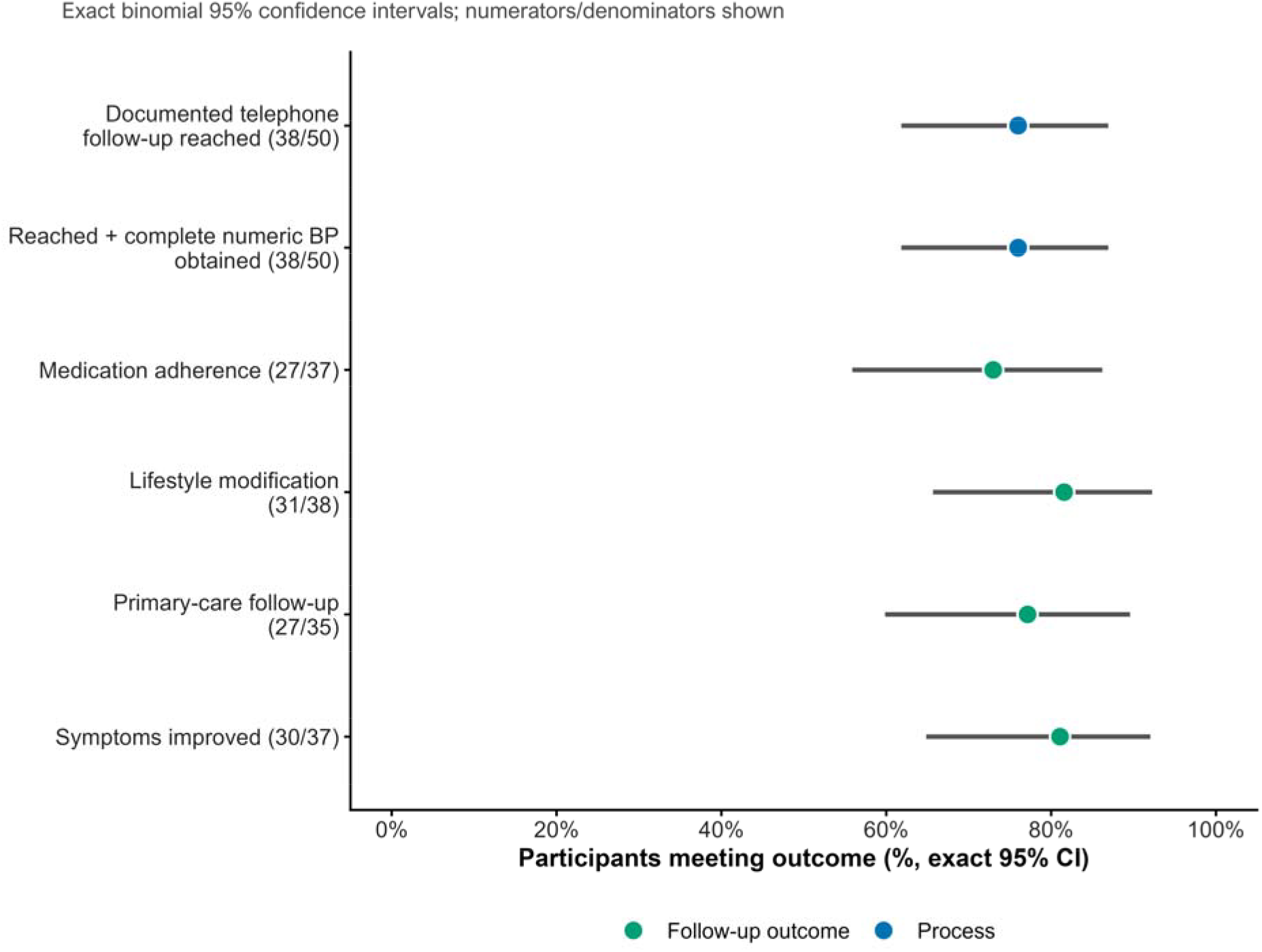
Feasibility, care engagement, and patient-reported follow-up outcomes. Points show observed proportions and horizontal lines show exact binomial 95% confidence intervals. Numerators and denominators are shown with each outcome. Behavioral outcomes use reached participants with nonmissing responses as the observed denominator.

## Discussion

### Principal findings

In this single-center QI initiative, structured hypertension education before ED discharge followed by telephone contact approximately 2 weeks later was associated with substantial short-term within-participant reductions in both SBP and DBP. More than half of participants with paired follow-up measurements met the strict <140/<90 mmHg control definition, and most participants achieved a clinically notable SBP reduction. Follow-up feasibility was moderate, with paired BP obtained from 76% of the enrolled cohort, while reported adherence, lifestyle change, primary-care attendance, and symptom improvement were frequent among reached participants with available responses.

### Interpretation in relation to existing evidence

Patient education and self-management interventions can improve BP, although average effects in controlled studies are generally more modest than the change observed here. A systematic review and meta-analysis by Foroumandi and colleagues reported mean reductions of approximately 5 mmHg in SBP and 4 mmHg in DBP with self-management education, and a later meta-analysis of individualized lifestyle education similarly found modest benefits. [9,10] The larger change observed in our project should therefore not be interpreted as evidence that this intervention is more efficacious. Participants entered with markedly elevated BP, and the uncontrolled pre-post design is particularly vulnerable to regression to the mean, medication changes after the ED encounter, differences in measurement conditions, and other time-varying factors.

Telephone-supported hypertension management has produced mixed results. Bosworth and colleagues demonstrated improved hypertension control with self-management strategies that included telephone support, and patient-tailored nurse telephone interventions have been developed to improve adherence and health behaviors.[11,12] In contrast, the MyHEART randomized clinical trial did not demonstrate a significant BP benefit from telephone coaching compared with usual care, despite improvements in some self-management behaviors.[13] These data support viewing telephone follow-up as a component of a broader care-transition strategy rather than as an isolated causal mechanism.

The ED is an important setting for such transition work because elevated BP is common in acute care, yet the evidence base for management and follow-up after acute-care detection remains less developed than in outpatient practice.[6] Our intervention attempted to bridge this gap with a simple workflow: standardized education at discharge, reinforcement of self-management, and early telephone contact. This approach is consistent with the WHO HEARTS emphasis on lifestyle counseling, standardized care processes, team-based approaches, and monitoring systems.[14] QI work in India has also shown that systematic process changes can improve hypertension follow-up, while longitudinal evidence from rural China supports regular follow-up as a component of BP management. [7,8]

The Pakistan context is relevant. Prior local literature has documented a substantial burden of uncontrolled hypertension and challenges with treatment, adherence, and risk-factor modification. [3-5] A low-cost intervention that strengthens continuity after an ED encounter may therefore be operationally attractive, particularly where outpatient access is inconsistent. However, implementation value should be judged not only by short-term BP change but also by reach, sustainability, workload, equity, and longer-term linkage to primary care.

### Strengths and limitations

Strengths include consecutive enrollment in a real-world ED, a simple intervention that could be incorporated into routine discharge workflows, patient-level paired analyses, explicit reporting of uncertainty, robust nonparametric and permutation analyses, a final data-integrity audit, and multiple sensitivity analyses for missing follow-up BP. The analysis also distinguishes observed patient-reported outcomes from conservative all-enrolled lower bounds and assesses baseline imbalance between participants with and without paired data.

Several limitations are important. First, the project had no concurrent control group; causal attribution is therefore not possible. Regression to the mean is a major concern because enrollment occurred in the setting of uncontrolled or elevated BP. Second, follow-up occurred over only approximately 2 weeks, so persistence of BP reduction and behavior change is unknown. Third, follow-up BP measurements could come from home monitors, clinics, or hospitals, introducing heterogeneity in devices, measurement technique, timing, and context. Fourth, medication initiation or intensification after the ED visit was not captured in sufficient detail to separate educational and treatment effects. Fifth, 12/50 participants did not contribute paired follow-up BP; baseline SMDs showed meaningful imbalance, particularly for sex, creating potential selection bias. Multiple-imputation and MNAR sensitivity analyses address assumptions about missing outcomes but cannot remove this bias.

Sixth, patient-reported adherence, lifestyle change, primary-care attendance, and symptom improvement were descriptive and subject to recall and social-desirability bias, with small amounts of item-level missingness. Seventh, the project was conducted at a single center with a small sample, limiting precision, subgroup analysis, and generalizability. Eighth, the strict <140/<90 mmHg threshold was a pragmatic project endpoint rather than an individualized treatment target; contemporary guideline-based targets depend on clinical context and patient characteristics.[1] Finally, the 10-mmHg SBP and 5-mmHg DBP responder thresholds were supportive descriptive analyses and should not be interpreted as prespecified causal treatment thresholds.

### Implications for future improvement cycles

Future PDSA cycles should prioritize improving follow-up reach and standardizing BP measurement. Practical steps include confirming multiple contact methods before discharge, scheduling the follow-up call before the patient leaves the ED, using reminder messages when feasible, documenting medication changes, and linking telephone follow-up directly to primary-care appointments. A subsequent evaluation should use standardized BP measurement, longer follow-up, repeated time points, and ideally a concurrent comparison group or staggered implementation design.

## Conclusion

Structured hypertension education at ED discharge followed by early telephone follow-up was associated with substantial short-term reductions in SBP and DBP among participants with paired measurements, and more than half met a strict <140/<90 mmHg BP-control definition. The intervention also achieved 76% follow-up with frequent reported engagement in medication adherence, lifestyle modification, and primary care. These findings support the feasibility of a low-cost transition-of-care strategy but do not establish a causal treatment effect. Controlled studies with standardized BP measurement and longer follow-up are needed before effectiveness can be inferred.

## Supporting information

SQUIRE checklist

## Data Availability

All data produced in the present work are contained in the manuscript.

## Declarations

### Ethics approval and consent to participate

This quality improvement project was approved by the Technical and Ethical Review Committee (TERC), a review wing of the Institutional Review and Research Advisory Board (IRRAB), Shaikh Zayed Medical Complex, Lahore, Pakistan (TERC ID: TERC/SC/INT/2026/111; approval date: 28 July 2026). Verbal informed consent was obtained from all participants before enrollment.

### Data and code availability

The statistical analysis code and aggregate analysis outputs have been retained by the study team. De-identified participant-level data are not publicly available, and any access would be subject to applicable institutional and ethics requirements.

## References

1. World Health Organization. Guideline for the pharmacological treatment of hypertension in adults. Geneva: World Health Organization; 2021. https://www.who.int/publications/i/item/9789240033986

2. Charchar FJ, Prestes PR, Mills C, Ching SM, Neupane D, Marques FZ, et al. Lifestyle management of hypertension: International Society of Hypertension position paper endorsed by the World Hypertension League and European Society of Hypertension. J Hypertens. 2024;42(1):23–49. doi:10.1097/HJH.0000000000003563.

3. Saleem F, Hassali AA, Shafie AA. Hypertension in Pakistan: time to take some serious action. Br J Gen Pract. 2010;60(575):449–450. doi:10.3399/bjgp10X502182.

4. Awan MUM, Akram Z, Mushtaq HH, Niazi HS, Irshad L, Imtiaz R, et al. Prevalence of hypertension in people aged 40 years and above. J Health Rehabil Res. 2024;4(2):1524–1529. doi:10.61919/jhrr.v4i2.1167.

5. Khan SA, Hafeez A, Zaka A, Khan SA, Ahmed A, Pervaiz F, et al. A randomized controlled trial of blood pressure reduction based on Disease Control Priorities 3 in Pakistan to manage and control hypertension. High Blood Press Cardiovasc Prev. 2023;30(4):357–366. doi:10.1007/s40292-023-00589-y.

6. Bress AP, Anderson TS, Flack JM, Ghazi L, Hall ME, Laffer CL, et al. The management of elevated blood pressure in the acute care setting: a scientific statement from the American Heart Association. Hypertension. 2024;81(8):e94–e106. doi:10.1161/HYP.0000000000000238.

7. Bharadwaj R, Kaviprawin M, Neelkanth N, Navkar VK, Azarudeen MJ, Parasuraman G, et al. Improving follow-up visits among individuals with hypertension: quality improvement project in the District Hospital, Seoni, Madhya Pradesh, India, 2021-2022. BMJ Open Qual. 2025;14(1):e003124. doi:10.1136/bmjoq-2024-003124.

8. Chen F, Yang E, Qing H, Wei Y, Tang S. The effect of follow-up on the blood pressure control: a longitudinal study in rural areas of China. Popul Health Metr. 2025;23(1):40. doi:10.1186/s12963-025-00406-9.

9. Foroumandi E, Kheirouri S, Alizadeh M. The potency of education programs for management of blood pressure through increasing self-efficacy of hypertensive patients: a systematic review and meta-analysis. Patient Educ Couns. 2020;103(3):451–461. doi: 10.1016/j.pec.2019.09.018.

10. Soltani D, Azizi B, Behnoush AH, Meysamie A, Aein A, Nayebirad S, et al. Is lifestyle modification with individual face-to-face education and counseling more effective than usual care for controlling hypertension? A systematic review and meta-analysis of randomized controlled trials. Health Educ Res. 2023;38(5):490–512. doi:10.1093/her/cyad028.

11. Bosworth HB, Olsen MK, Grubber JM, Neary AM, Orr MM, Powers BJ, et al. Two self-management interventions to improve hypertension control: a randomized trial. Ann Intern Med. 2009;151(10):687–695. doi:10.7326/0003-4819-151-10-200911170-00148.

12. Bosworth HB, Olsen MK, Gentry P, Orr M, Dudley T, McCant F, et al. Nurse administered telephone intervention for blood pressure control: a patient-tailored multifactorial intervention. Patient Educ Couns. 2005;57(1):5–14. doi: 10.1016/j.pec.2004.03.011.

13. Hoppe KK, Smith M, Birstler J, Kim K, Sullivan-Vedder L, LaMantia JN, et al. Effect of a telephone health coaching intervention on hypertension control in young adults: the MyHEART randomized clinical trial. JAMA Netw Open. 2023;6(2):e2255618. doi:10.1001/jamanetworkopen.2022.55618.

14. World Health Organization. HEARTS: technical package for cardiovascular disease management in primary health care. Geneva: World Health Organization; 2018. https://www.who.int/publications/i/item/WHO-NMH-NVI-18-14

15. Ogrinc G, Davies L, Goodman D, Batalden P, Davidoff F, Stevens D. SQUIRE 2.0 (Standards for QUality Improvement Reporting Excellence): revised publication guidelines from a detailed consensus process. BMJ Qual Saf. 2016;25(12):986–992. doi:10.1136/bmjqs-2015-004411.

