## Supplemental Document for "Structured Hypertension Education and Telephone Follow-up After Emergency Department Discharge: A Prospective Quality Improvement Initiative in Pakistan"

**Supplementary Material**

### Supplementary Methods

#### Final analysis population and data audit

The final locked analytic dataset contained 50 rows and 50 unique patient identifiers, with no duplicate identifiers. Thirty-eight participants were explicitly documented as reached by telephone and had both numeric follow-up SBP and DBP; these participants formed the primary paired cohort. No participant coded as reached lacked a complete BP pair, and no complete BP pair was present without documented reach. There were no partial SBP/DBP pairs.

Analytic BP-control, BP-improvement, and BP-reduction variables were recalculated from the numeric baseline and follow-up measurements rather than taken from manually entered summary flags. The final audit identified no disagreements between the workbook flags and the recalculated values, and the exported final audit problem-row file was empty.

#### Primary and robustness analyses

The primary estimand was mean within-participant SBP reduction, defined as baseline SBP minus follow-up SBP, among participants with complete paired measurements. DBP reduction was analyzed analogously. Nonparametric bootstrap 95% CIs used 10,000 resamples. Paired t tests, Wilcoxon signed-rank tests, 20% trimmed means with bootstrap CIs, and 50,000-draw sign-flip permutation tests were used to assess robustness to distributional assumptions. Cohen dz and small-sample corrected Hedges gz summarized standardized paired effect size.

Strict BP control was defined as follow-up SBP <140 mmHg and DBP <90 mmHg. An inclusive <=140/<=90 mmHg definition was examined as a threshold sensitivity analysis. Supportive responder thresholds were an SBP reduction >=10 mmHg and DBP reduction >=5 mmHg. Exact binomial 95% CIs were used for proportions.

Baseline differences between participants in and outside the paired cohort were summarized using standardized mean differences for age, sex, baseline SBP, and baseline DBP. These are descriptive measures of potential selection or attrition bias and were not treated as hypothesis tests.

#### Missing-data sensitivity analyses

Four complementary missing-data approaches were used. First, a conservative no-change analysis carried baseline BP forward for all participants outside the primary paired cohort. Second, multiple imputation under a missing-at-random assumption used predictive mean matching with 50 imputations and 30 iterations; baseline SBP, baseline DBP, age, and sex were used as predictors of missing follow-up BP. Scalar estimates were pooled using Rubin rules.

Third, delta-adjusted missing-not-at-random analyses examined prespecified adverse increments of +5, +10, +20, and +30 mmHg for imputed follow-up SBP and +2.5, +5, +10, and +15 mmHg for imputed follow-up DBP. Positive deltas represent systematically worse follow-up BP among participants with missing measurements. Selected moderate and severe scenarios (+10/+5 mmHg and +20/+10 mmHg for SBP/DBP, respectively) are presented in Supplementary Table S1. Fourth, deterministic sensitivity analyses assigned progressively worse average BP changes to the 12 participants outside the primary cohort and calculated the mean all-enrolled reduction. Tipping points quantified the mean worsening among the 12 nonprimary participants that would be required to reduce the overall mean change to zero.

### Supplementary Tables

#### Supplementary Table S1. Missing-data robustness analyses

| **Endpoint** | **Analysis** | **Mean reduction, mmHg** | **95% CI** |
| --- | --- | --- | --- |
| SBP | Primary: reached + complete numeric BP | 31.8 | 24.4 to 39.3 |
| DBP | Primary: reached + complete numeric BP | 10.4 | 6.7 to 13.9 |
| SBP | All enrolled: nonprimary assumed no change | 24.1 | 17.6 to 30.8 |
| DBP | All enrolled: nonprimary assumed no change | 7.9 | 4.9 to 11.0 |
| SBP | MI under MAR | 32.6 | 26.2 to 39.1 |
| DBP | MI under MAR | 10.2 | 6.7 to 13.7 |
| SBP | MI MNAR: missing follow-up +10 SBP / +5 DBP | 30.2 | 23.8 to 36.7 |
| DBP | MI MNAR: missing follow-up +10 SBP / +5 DBP | 9.0 | 5.4 to 12.6 |
| SBP | MI MNAR: missing follow-up +20 SBP / +10 DBP | 27.8 | 21.1 to 34.6 |
| DBP | MI MNAR: missing follow-up +20 SBP / +10 DBP | 7.8 | 4.0 to 11.6 |

Positive estimates indicate lower follow-up BP. Bootstrap CIs are used for observed/no-change analyses; Rubin-pooled CIs are used for multiple-imputation analyses.

#### Supplementary Table S2. Deterministic missing-data sensitivity and tipping points

| **Endpoint** | **Assumed mean change among 12 nonprimary participants, mmHg** | **All-enrolled mean reduction, mmHg** |
| --- | --- | --- |
| SBP | 0 | 24.1 |
| SBP | -5 | 22.9 |
| SBP | -10 | 21.7 |
| SBP | -20 | 19.3 |
| SBP | -30 | 16.9 |
| SBP | -40 | 14.5 |
| DBP | 0 | 7.9 |
| DBP | -2.5 | 7.3 |
| DBP | -5 | 6.7 |
| DBP | -10 | 5.5 |
| DBP | -15 | 4.3 |
| DBP | -20 | 3.1 |

Tipping point: to make the all-enrolled mean change equal to zero, the 12 participants outside the primary cohort would need an average SBP reduction of -100.6 mmHg and DBP reduction of -32.9 mmHg; negative reductions denote worsening.

#### Supplementary Table S3. Distribution of individual BP change

| **Endpoint** | **Direction** | **n** | **%** |
| --- | --- | --- | --- |
| DBP | Improved | 26 | 68.4 |
| DBP | No change | 9 | 23.7 |
| DBP | Worsened | 3 | 7.9 |
| SBP | Improved | 35 | 92.1 |
| SBP | No change | 1 | 2.6 |
| SBP | Worsened | 2 | 5.3 |

#### Supplementary Table S4. Supplementary hemodynamic outcomes

| **Outcome** | **N** | **Baseline mean (SD)** | **Follow-up mean (SD)** | **Mean reduction** | **95% bootstrap CI** | **P (paired t)** |
| --- | --- | --- | --- | --- | --- | --- |
| Pulse pressure | 38 | 69.7 (16.6) | 48.3 (9.8) | 21.4 | 15.6 to 27.1 | <0.001 |
| Mean arterial pressure | 38 | 118.0 (11.1) | 100.4 (8.2) | 17.5 | 13.1 to 21.9 | <0.001 |

#### Supplementary Table S5. Conservative all-enrolled lower bounds for patient-reported outcomes

| **Outcome** | **Events/N** | **Lower-bound %** | **Exact 95% CI** |
| --- | --- | --- | --- |
| Medication adherence | 27/50 | 54.0 | 39.3 to 68.2 |
| Lifestyle modification | 31/50 | 62.0 | 47.2 to 75.3 |
| Primary-care follow-up | 27/50 | 54.0 | 39.3 to 68.2 |
| Symptoms improved | 30/50 | 60.0 | 45.2 to 73.6 |

These are conservative lower-bound sensitivity values using all 50 enrolled participants as the denominator; participants not reached or with missing responses are not interpreted as observed failures.

### Supplementary Figures

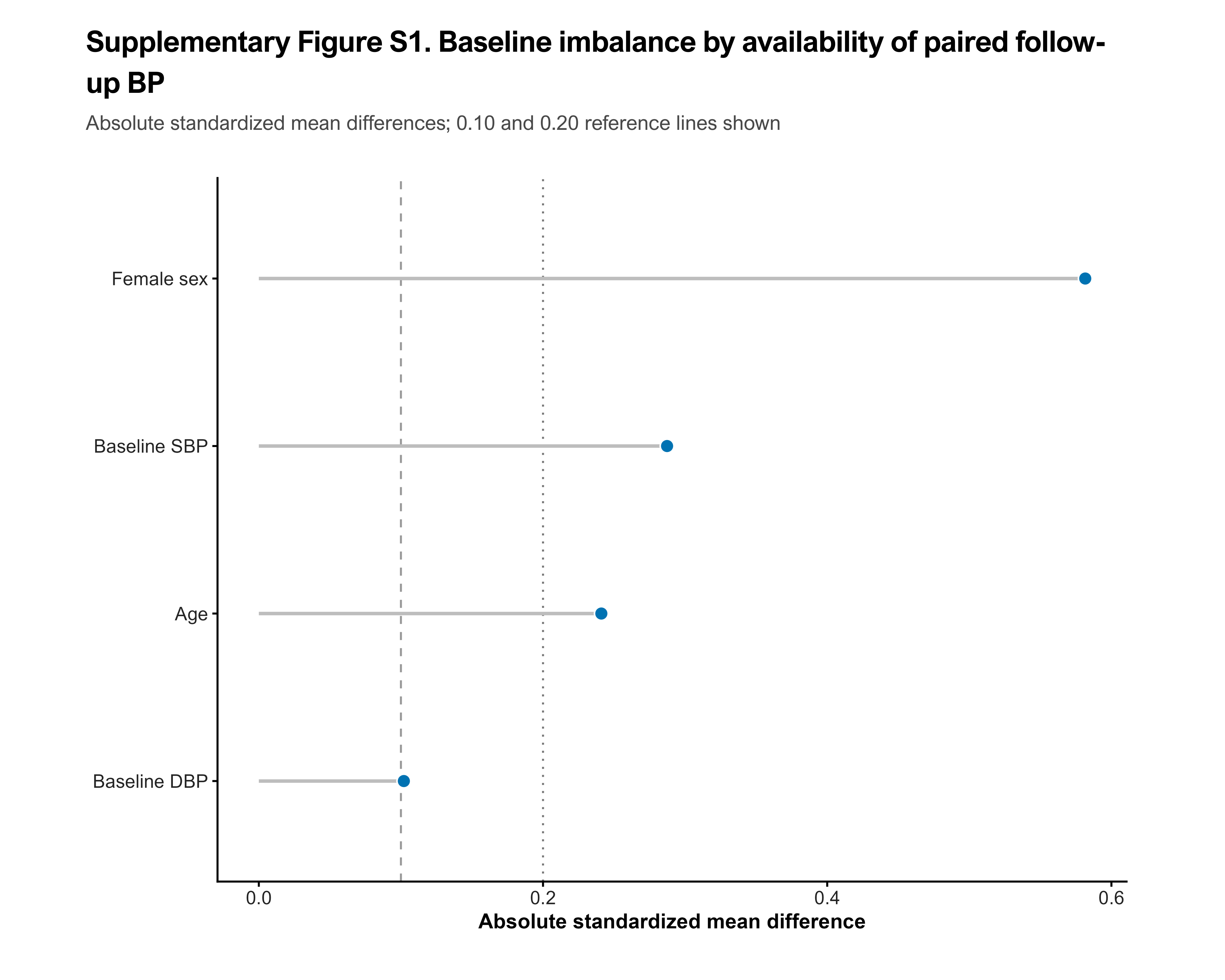

Supplementary Figure S1. Baseline imbalance between participants in the final paired BP cohort and those without paired follow-up BP. Absolute standardized mean differences are shown for the baseline variables available in the final Cleaned_Data sheet: age, sex, baseline SBP, and baseline DBP.

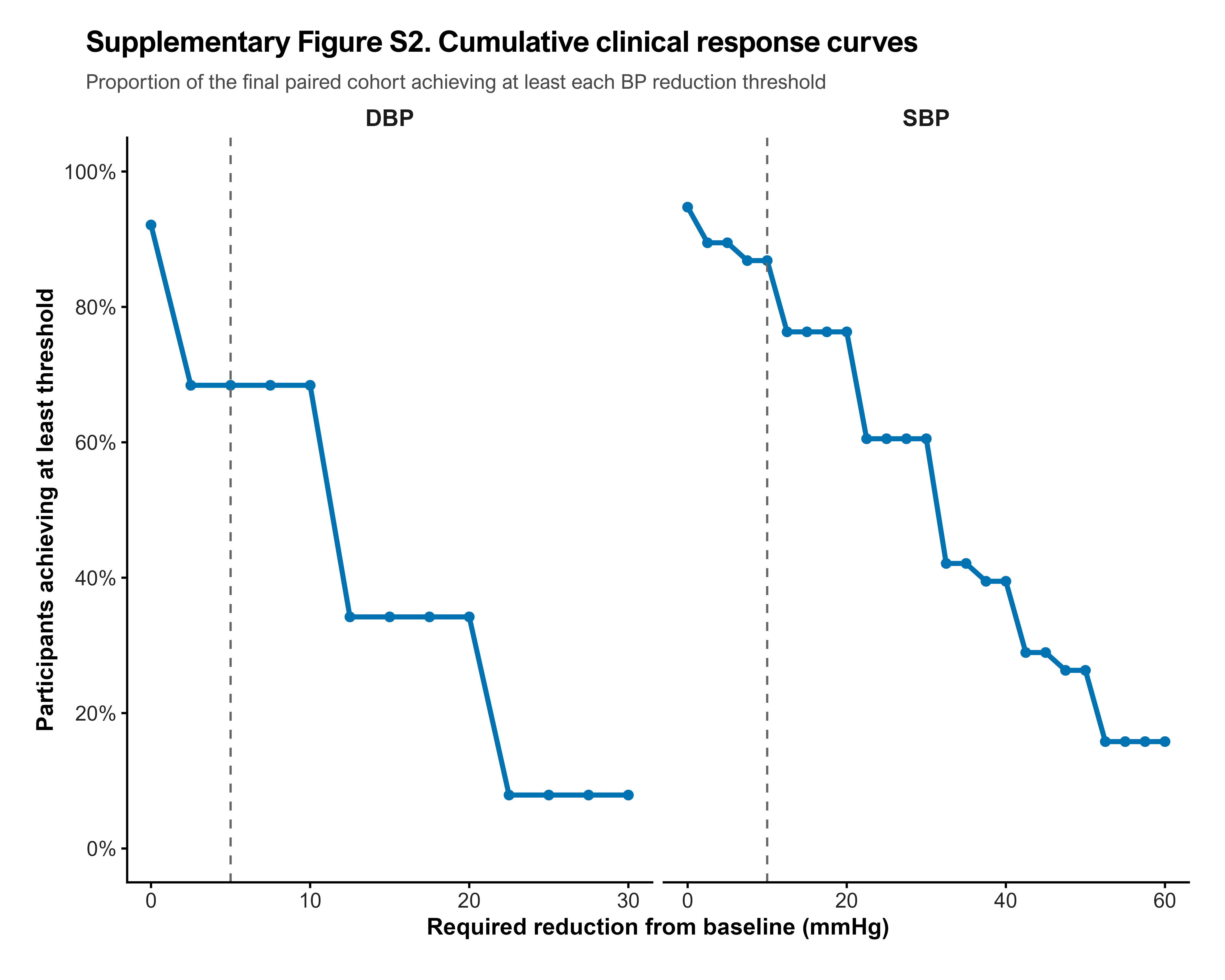

Supplementary Figure S2. Cumulative clinical response curves. Curves show the proportion of the final primary cohort achieving at least each specified SBP or DBP reduction threshold. Dashed vertical lines mark the supportive responder thresholds used in the manuscript.

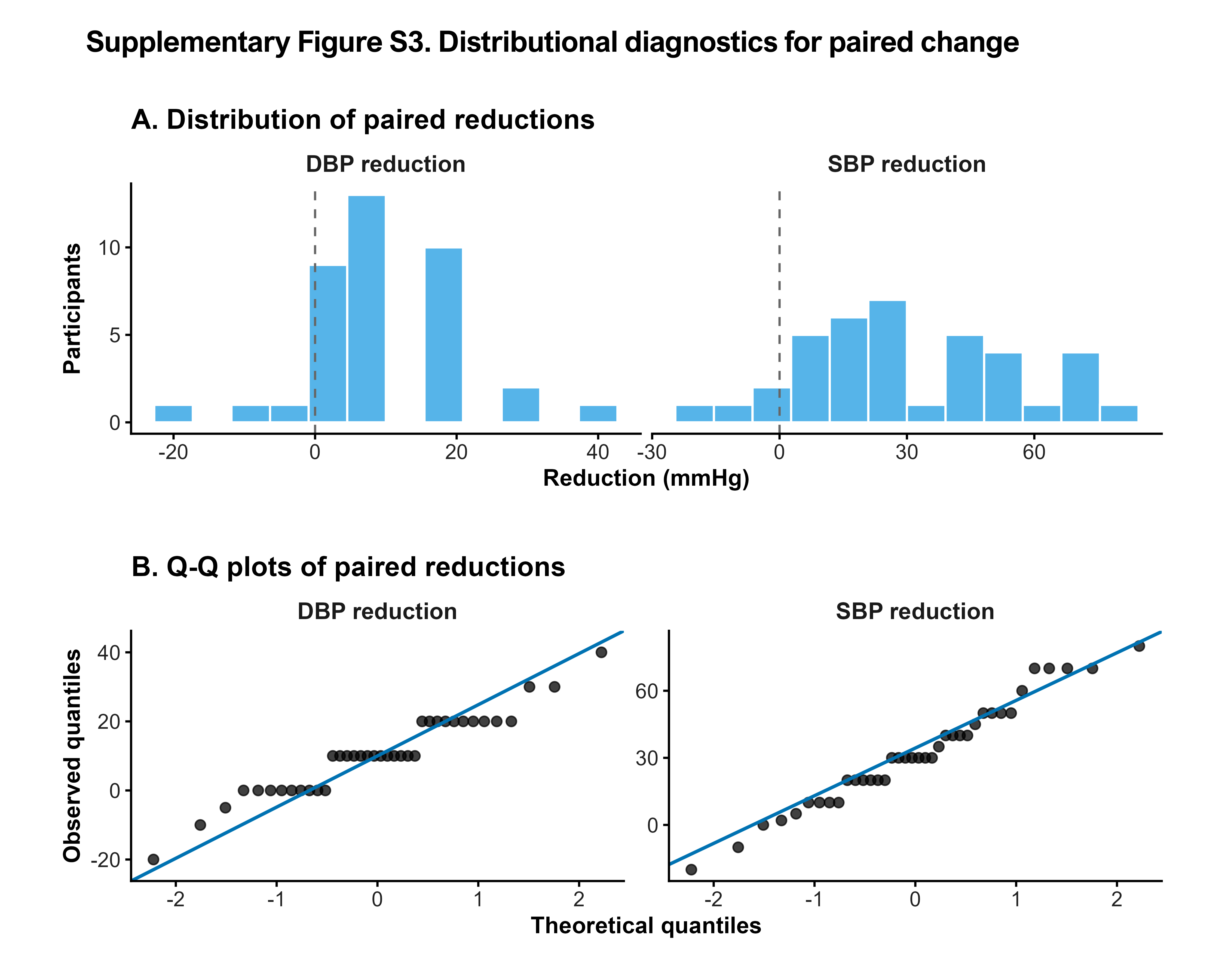

Supplementary Figure S3. Distributional diagnostics for paired blood pressure change. Histograms and Q-Q plots display observed paired SBP and DBP reductions. Primary estimation is accompanied by bootstrap, Wilcoxon, trimmed-mean, and sign-flip permutation sensitivity analyses.
