## Supplementary material for "Structured Hypertension Education and Telephone Follow-up After Emergency Department Discharge: A Prospective Quality Improvement Initiative in Pakistan": SQUIRE checklist

**SQUIRE 2.0 Checklist**

Reporting guideline: Ogrinc G, Davies L, Goodman D, et al. SQUIRE 2.0. BMJ Qual Saf. 2016;25:986-992. doi:10.1136/bmjqs-2015-004411.

| **Item** | **SQUIRE 2.0 topic** | **Checklist requirement** | **Where addressed in manuscript** | **Page(s)** |
| --- | --- | --- | --- | --- |
| 1 | Title | Indicate that the manuscript concerns an initiative to improve healthcare and identify the principal intervention/topic. | Title identifies structured hypertension education and telephone follow-up as a prospective quality improvement initiative. | p. 1 |
| 2 | Abstract | Summarize key information from all sections using the journal-specified abstract format. | Structured abstract reports background, setting/design, intervention, primary analysis, principal results, limitations, and conclusion. | pp. 2–3 |
| 3 | Problem description | Describe the nature and significance of the local problem. | Introduction describes uncontrolled hypertension and weak continuity after ED discharge, with relevance to Pakistan and resource-constrained settings. | p. 3 |
| 4 | Available knowledge | Summarize what is currently known about the problem and relevant interventions. | Introduction reviews hypertension management, acute-care transition needs, follow-up QI work, and longitudinal follow-up evidence. | p. 3 |
| 5 | Rationale | Describe the informal or formal rationale/theory explaining why the intervention was expected to work. | Rationale links standardized education and early telephone reinforcement to self-management, BP reassessment, and linkage to longitudinal care; PDSA framework is described. | pp. 3–4 |
| 6 | Specific aims | State the purpose of the project and the specific aims. | End of Introduction specifies primary aim (short-term within-participant SBP change) and secondary aims (DBP, control/responder outcomes, feasibility, and patient-reported care behaviors). | p. 3 |
| 7 | Context | Describe contextual elements considered important to the intervention. | Methods identifies a single ED at Shaikh Zayed Hospital, Lahore, project period, resource context, eligibility, and local continuity/education gap. | pp. 3–4 |
| 8 | Intervention(s) | Describe the intervention in sufficient detail for replication; describe the team involved and changes over time when relevant. | Methods describes the standardized discharge education, the structured approximately 2-week telephone follow-up, the staff roles responsible for intervention delivery, and the initial PDSA cycle. No intervention modifications occurred during the reported project period. | p. 4 |
| 9 | Study of the intervention(s) | Describe the approach used to assess whether observed outcomes were due to the intervention and how the approach evolved. | Methods identifies the prospective pre-post QI design and describes paired estimation, attrition assessment, and multiple missing-data sensitivity analyses; the Discussion explicitly limits causal inference because no concurrent control group was used. | pp. 3–5; pp. 8–10 |
| 10 | Measures | Describe process and outcome measures, rationale, definitions, validity/reliability considerations, and contextual measures. | Outcome Measures defines SBP/DBP change, strict and inclusive control thresholds, responder outcomes, follow-up reach, and patient-reported outcomes. BP source heterogeneity is explicitly acknowledged. | pp. 4–5; p. 9 |
| 11 | Analysis | Describe qualitative and quantitative methods used to draw inferences, including methods for variation and time effects when applicable. | Data integrity and statistical analysis describes bootstrap CIs, paired t tests, Wilcoxon tests, permutation tests, effect sizes, exact binomial CIs, SMDs, BOCF/no-change sensitivity, multiple imputation, MNAR deltas, and tipping-point analysis. | p. 5 |
| 12 | Ethical considerations | Describe ethical aspects of implementing and studying the intervention and how they were addressed. | Methods and Declarations report approval by the Technical and Ethical Review Committee (TERC), a review wing of the Institutional Review and Research Advisory Board (IRRAB), Shaikh Zayed Medical Complex, Lahore, Pakistan (TERC ID: TERC/SC/INT/2026/111), together with the consent procedure. | pp. 5–6; p. 10 |
| 13 | Results | Report intervention steps, evolution, process measures, outcome measures, contextual elements, associations, unintended consequences, and missing data. | Results reports participant reach, baseline and attrition imbalance, paired blood-pressure estimates with confidence intervals, BP-control and responder outcomes, process and patient-reported outcomes, and missing-data robustness analyses. Unintended consequences were not specifically reported as an outcome. | pp. 6–7; Tables pp. 11–13; Figures pp. 16–20 |
| 14 | Summary | Summarize key findings, including relevance to rationale and aims. | Discussion opens with the principal findings in relation to the project aims. | pp. 7–8 |
| 15 | Interpretation | Interpret associations between intervention and outcomes, compare with literature, discuss impact of context, costs/trade-offs where relevant, and reasons for differences. | Discussion compares results with education and telephone-support literature, emphasizes the uncontrolled design, explains likely contributors to the large observed change, and discusses Pakistan/resource-constrained context and implementation value. Formal cost data were not collected. | pp. 8–9 |
| 16 | Limitations | Discuss limits to generalizability, internal validity, imprecision, and efforts to minimize/adjust for limitations. | Strengths and limitations addresses lack of control group, regression to the mean, short follow-up, heterogeneous BP measurement, medication changes, missing paired data/selection bias, self-report, sample size, single-center design, and endpoint definitions; sensitivity analyses are described. | pp. 9–10 |
| 17 | Conclusions | Describe usefulness, sustainability, spread, implications for practice and further study, and suggested next steps. | Conclusion and future improvement-cycle section describe feasibility while avoiding causal claims and recommend improved follow-up reach, standardized BP measurement, longer follow-up, medication documentation, and a concurrent comparison design. | p. 10 |
| 18 | Funding | Describe funding sources and the role of funders, if any. | No external or internal funding was received for this quality improvement project. | p. 10 |
